# Serum Vitamin D Levels Are Not Associated with Obstructive Lung Disease in the General Population: An NHANES Analysis (2007-2008 to 2009-2010)

**DOI:** 10.1101/2020.11.10.20229310

**Authors:** Mohamed I Seedahmed, Aaron D Baugh, Jordan A Kempker

**Affiliations:** Department of Medicine, Division of Pulmonary, Allergy, Critical Care and Sleep Medicine, University of California San Francisco, California, USA; University of California San Francisco Helen Diller Medical Center, San Francisco, California, USA; San Francisco Veterans Affairs Medical Center, San Francisco, California, USA; Grady Memorial Hospital, Atlanta, Georgia, USA; Department of Medicine, Division of Pulmonary, Allergy, Critical Care and Sleep Medicine, Emory University, Atlanta, Georgia, USA

**Author notes:** **Corresponding Author: Mohamed I Seedahmed, MD, MPH**, Division of Pulmonary, Critical care, allergy and Immunology, and Sleep., Department of Medicine, University of California, San Francisco, University of California San Francisco Helen Diller Medical Center, 513 Parnassus Ave., HSE 1314, Box 0111, San Francisco, CA 94143. Email Addresses : MIS, ADB, JAK. **Authors’ Contributions:** Conceived and designed the study research: MIS, JAK. Developed study protocol: MIS, JAK. Worked on the methods: MIS, JAK. Analyzed and Interpreted data: MIS, ADB, JAK. Prepared the manuscript: MIS, ADB, JAK.

**Keywords:** Obstructive lung disease, serum 25(OH)D, total Vitamin D, healthcare disparity, spirometry, the National Health and Nutrition Examination Survey (NHANES)

## Abstract

**Background:** Obstructive lung disease is a significant cause of morbidity and healthcare burden within the United States. A growing body of evidence has suggested that vitamin D levels can influence the course or incidence of obstructive lung disease. However, there is an insufficient previous investigation of this association.

**Study Design and Methods:** We used the National Health and Nutrition Examination Survey (NHANES) cycles 2007-2008 and 2009-2010 spirometry results of individuals aged 40 years and older to assess the association between serum 25(OH)D levels and obstructive lung disease, as defined by the American Thoracic Society using the lower limit of normal (LLN). We used stage multivariate survey-logistic regression with backward selection.

**Results:** The final model included body mass index, pack-years smoking history, and ethnicity. In the primary model, there was no association between vitamin D levels and obstructive lung disease. We noted an association between “Other Hispanic” self-identified race and serum Vitamin D levels wherein higher levels were associated with higher odds of obstructive lung disease in this ethnicity, but not among other racial or ethnic groups (OR= 1.48, p= 0.02).

**Conclusions:** Serum Vitamin D levels among adults are not associated with the odds of obstructive lung disease in the general population. Results among non-Mexican Hispanic participants highlight the need for further research in minority populations. More work is needed to address the course and incidence of lung disease in the United States.

**RESEARCH IN CONTEXT:** *What is the key question?:* In the general population, is there an independent association between Vitamin D and obstructive lung disease after controlling for relevant covariates?

*What is the bottom line?:* In exploring whether serum vitamin D levels are associated with odds of obstructive lung disease in the general US population, we did not find an independent association in the overall sample.

*Why read on?:* This paper adds nuance to the broad understanding of vitamin D’s role in lung pathophysiology.

## INTRODUCTION

Obstructive airway disease is a significant public health concern in the United States. COPD was estimated to cost 16.4 million lost workdays in 2010 and has greater odds of prompting an end to employment than diabetes or heart disease.[1,2] In the next 20 years, asthma is projected to cost over $900 billion to the US economy.[3] Together with COPD, it is the fourth leading cause of death in the United States.[4] Given the enormity of these challenges, there is an urgent need to identify interventions that can reduce the incidence and burden of obstructive airway disease.

While vitamin D has been classically described regarding healthy bone metabolism, there has been increased attention towards its extra-skeletal physiologic actions in recent years. Low levels of 25-hydroxyvitamin D (25(OH)D) have been linked to clinical conditions such as rickets, hypertension, ischemic heart disease, diabetes mellitus type 1, some cancers, osteoporosis, and infections.[5] Its role in lung development and pathophysiology has also received increasing attention. Gestational deficiency is negatively associated with later pulmonary function, and confers increased odds of asthma.[6,7] In adults, vitamin D deficiency has been linked to an increased likelihood of respiratory diseases.[8] Besides, many of the demographic groups with a higher incidence of Vitamin D deficiency[9] also report more significant morbidity from obstructive airway diseases.[10] Data from the National Health and Nutrition Examination Survey (NHANES) 2005 to 2006 revealed that 41.6 percent of adults’ participants ≥20 years-old had vitamin D deficiency, with higher prevalence among non-Hispanic blacks (82.1%), those with no college education, and those with body-mass-index more than 30.[9] Also, data from NHANES 2001-2004 demonstrated that older age, female sex, winter season, and smoking are associated with vitamin D deficiency.[11]

However, much of the adult literature has been reported in diseased populations. In these populations, associations can be highly confounded. Vitamin D levels are influenced by sun exposure and, therefore, maybe a proxy for disability.[12] Further exploring the relationship between Vitamin D and pulmonary health in adults would benefit from extensive, robust studies of the general population to understand this association and its mediators. From there, the goal of this study was to utilize a large, representative sample of US adults to examine the relationship between vitamin D status and obstructive lung disease patterns among US adults aged 40 years and older.

## METHODS

### Data Source & Study Design

NHANES is a major program of the National Center for Health Statistics (NCHS) which is part of the Centers for Disease Control and Prevention (CDC). It is an ongoing national survey designed to assess the health of the general US population. Data are collected annually on a 2-year cycle, using a multistage, probability-sampling design to generate population-level estimates. The national survey employs a design variable in order to approximate the civilian, non-institutionalized US population. NHANES 2007-2010 included spirometry exams conducted according to the technical recommendations of the American Thoracic Society (ATS) for procedures and equipment.[13]We included all participants from the 2007-2008 and 2009-2010 survey cycles who were at least 40 years of age, completed spirometry, and had measured serum 25 (OH)D concentrations (Figure 1). Per their study protocols, NHANES excluded participants with BMI>40, a history of tuberculosis; supplemental oxygen usage; a history of hemoptysis or retinal detachment on attempted spirometry; or any recent stroke, active cardiovascular disease, retinal, thoracic, or abdominal surgery.

**Figure 1.**
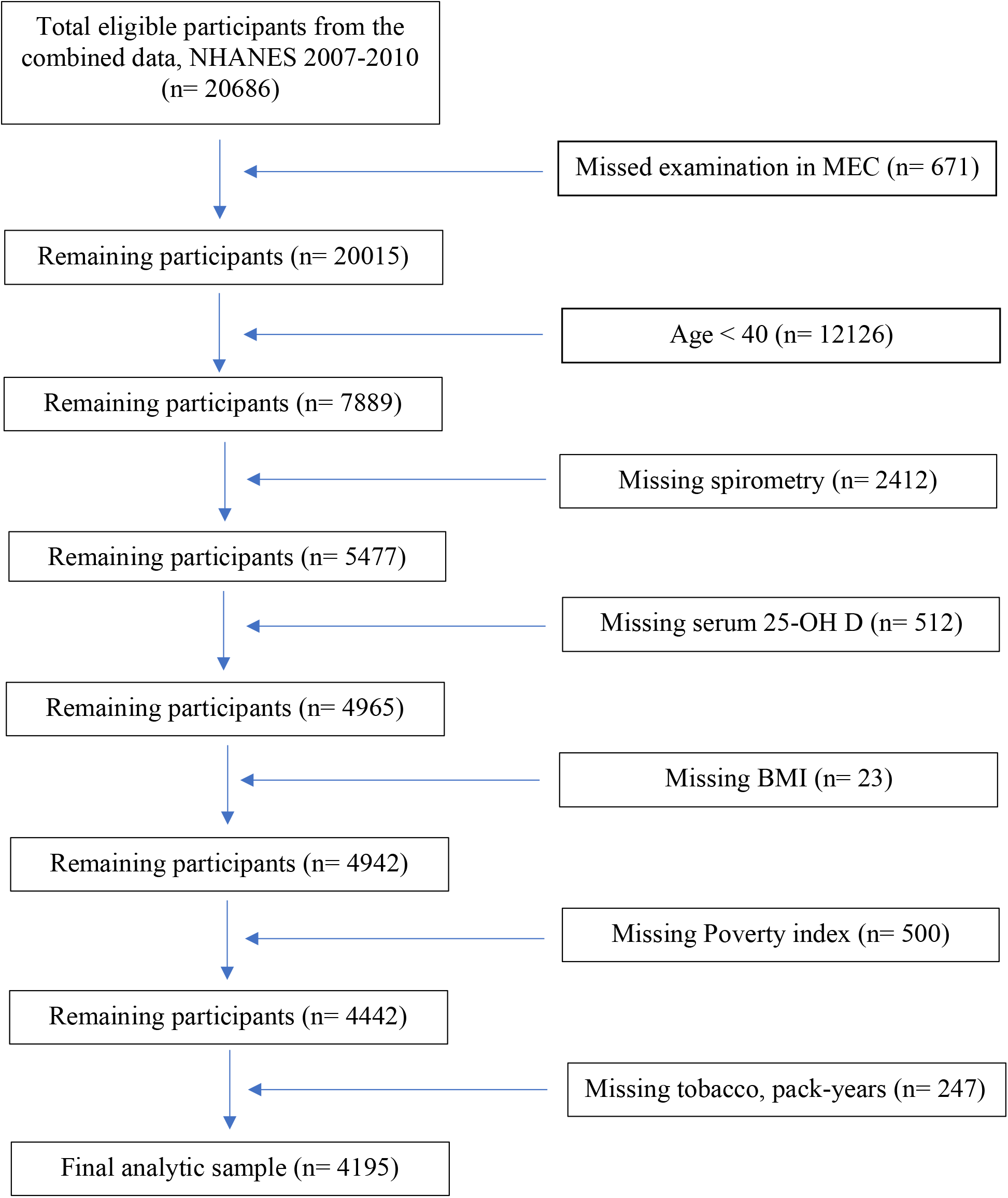
STROBE flow chart-sample selection criteria for the association between serum 25-hydroxyvitamin D concentration and baseline FEV1/FVC. Abbreviations-NHANES: National Survey of the National Center for Health Statistics; MEC: Mobile Examination Centers; 25-OH D: Total Vitamin D; BMI: Body Mass Index.

Public-use data from NHANES were obtained from files available on the NCHS website.[14] The data were sorted, merged, and concatenated using the unique sequence number given to each NHANES participants, in addition to a specific identifier number that code for the 2-year cycle, 2007-2008, and 2009-2010. This Study was approved by the NCHS Research Ethics Review Board (ERB).[15]

### Exposure Variable

The NHANES 2007-2010 used ultra-high-performance liquid chromatography-tandem mass spectrometry (LC-MS/MS) for all vitamin D measurements.[13] The primary exposure of interest, serum 25(OH)D concentration, was defined as low if the concentration was <30 nmol/L and adequate if the concentration was ≥ 30 nmol/L. Nevertheless, because the optimal 25(OH)D concentration for the normal respiratory function is uncertain, we also explored 25(OH)D as a continuous variable and as an ordinal variable based on recommendations from the Institute of Medicine into a 3-category variable: deficiency (<30 nmol/L), normal level (30 - 75 nmol/L), and above normal level (>75 nmol/L).[16] For the final analyses, we created a new continuous vitamin D variable, divided by 25, for which a change in 1 unit will equal a change in 25 nmol/L of vitamin D.

### Outcome Variable

The primary outcome variable in our analysis was the presence or absence of obstructive lung disease. We employed the ATS definition of obstructive disease as having a ratio of forced expiratory volume in the 1^st^ second to forced vital capacity (FEV_1_/FVC), which is less than the 5^th^ percentile lower limit of normal (LLN) observed in the healthy, non-smoking population.[17],[18] Per contemporaneous ATS guidelines, the spirometric measurements were re-coded to calculate the LLN using reference equations developed in 1999 from participants in NHANES III.[17],[19]

### Additional Covariates

Smoking is recognized as the most crucial risk factor for the development of obstructive lung disease.[20] Low body mass index (BMI) is associated with increased COPD risk and mortality.[21] Both BMI and pack-years smoking history were measured as continuous variables. Because seasonality can affect performance on spirometry[22], we included a 6-month interval binary variable to control for the time of a participant’s testing. Age, gender, race, and income have important associations with vitamin D level in the United States and were all measured as covariates.[23] Race/ethnicity was reported categorically according to the US Census classifications. Income was measured using the income-to-poverty ratio (IPR) calculated as a ratio of the reported household income to the national poverty threshold of the given year reported by the US Census Bureau.[24]

### Analyses

All statistical analyses were performed with SAS software (SAS, version 9.4, SAS Institute, Inc, Cary, North Carolina). Descriptive statistics were computed. For all analyses, we utilized the NHANES 2007-2010 sample weights previously calculated for the combined two survey cycles considered.[25] The PROC SURVEY procedure was employed to account for the intricate sampling design of NHANES. We followed the Taylor Series linearization method and made an assumption of non-random missingness for variance estimation, per NHANES analytic guidelines.[26] We also used domain analysis to refine our variance estimate.

In our primary analysis, we developed a priori multivariate survey logistic regression model of the association between obstructive lung disease, defined as FEV1/FVC< LLN, and serum 25(OH)D concentration. Variables were pre-selected based on identifying major determinants of obstructive lung disease in the previous medical literature. Following a pre-specified multistep approach, multivariate models were adjusted for age, gender, race/ethnicity, smoking in pack-years, BMI, the season of examination, and PIR. The modeling building strategy started with assessing for collinearity.[27] Following this step, we assessed for interaction terms between serum 25 (OH)D and other covariates. Next, we conducted a confounding assessment. The prevalence odds ratio of the a priori model was compared to all confounders’ possible subsets using the 10% rule. Last, we performed a backward stepwise regression selection of the covariates to develop the final model, the cut-off for inclusion was *p <* 0.30. Our final model included race/ethnicity, smoking in pack-years, BMI, and IPR. For all model estimates, we defined significance as nonoverlapping 95% confidence intervals and *p 0*.*05*. Moreover, we performed a secondary subgroup analysis for the association between serum 25(OH)D and obstructive lung disease stratified by race/ethnicity. A planned tertiary analysis by the Global Initiative for Chronic Obstructive Lung Disease (GOLD) stages was not completed due to the small number of participants with Stage 3 and 4 disease.

To help understand the differences between each race/ethnicity subgroups among subjects with obstructive lung disease defined by LLN, we computed the mean of post-bronchodilator FEV1 (% predicted) and serum 25(OH)D for each race/ethnicity category. Additionally, we used the Ellipse statement to graphically plot the predicted ellipses for each race/ethnicity subgroups.

## RESULTS

Of the 20,686 participants from NHANES 2007-2010, 20,015 had conducted the survey and examinations in the mobile examination centers (MEC). 4,195 met the inclusion criteria for our study (Figure 1).

### Study Participants’ Characteristics as related to the Baseline FEV1/FVC

Fourteen and half of a percent (*n*= 777) of the study population met the diagnostic criteria for obstructive lung disease with a baseline FEV1/FVC<LLN (Table 1). They were more likely non-Hispanic Whites (61% vs. 47%, *p*=<.0001), greater than 10 pack-year total lifetime smokers (55% vs. 26%, *p*= <.0001), to have a higher mean of serum 25(OH)D concentration (71 ± 1.14, p=<.0001), and to have a higher age mean (56.1 vs. 54.7, *p*=<.0001). Compared with participants with FEV1/FVC ≥ LLN, they were more likely Hispanics (29% vs. 16%, *p*= <.0001), and likely to be obese with BMI ≥ 30 (42% vs. 27%, *p*=<0001). There were no significant differences between the two groups, FEV1/FVC below and above LLN, by gender, PIR, or seasonality of exam administration.

**Table 1.** Demographics and Clinical Characteristics of Study Participants by Baseline FEV1/FVC ratio below and above LLN a, NHANES, 2007-2010.

| Characteristic | Baseline FEV1/FVC<br>< LLN <sup>b</sup> |  | Baseline FEV1/FVC<br>≥ LLN <sup>b</sup> |  | p<sup>c</sup> |
| --- | --- | --- | --- | --- | --- |
|  | N | % | N | % |  |
|  | 777 | 14.5 | 4700 | 85.5 |  |
| Age, years |  |  |  |  |  |
| 40 – 49 | 195 | 25 | 1459 | 31 | <.0001 |
| 50 – 59 | 202 | 26 | 1270 | 27 |  |
| 60 – 69 | 210 | 27 | 1219 | 26 |  |
| ≥ 70 | 170 | 22 | 752 | 16 |  |
| Mean ± SE <sup>d</sup> | 56.1 ± 0.41 |  | 54.7 ± 0.22 |  | <.0001 |
| Race |  |  |  |  |  |
| Mexican American | 72 | 9 | 852 | 18 | <.0001 |
| Other Hispanic | 55 | 7 | 536 | 11 |  |
| Non-Hispanic White | 474 | 61 | 2230 | 47 |  |
| Non-Hispanic Black | 154 | 20 | 892 | 20 |  |
| Other race or Multi-racial | 22 | 3 | 190 | 4 |  |
| Gender |  |  |  |  |  |
| Male | 445 | 57.3 | 2280 | 48.5 | 0.21 |
| Female | 332 | 42.7 | 2420 | 51.5 |  |
| BMI <sup>e</sup>, kg/m<sup>2</sup> |  |  |  |  |  |
| < 18.5 | 26 | 3 | 33 | 1 | <.0001 |
| 18.5 – 25 | 258 | 33 | 946 | 20 |  |
| 25 – 30 | 283 | 37 | 1716 | 37 |  |
| ≥ 30 | 205 | 27 | 1987 | 42 |  |
| Missing <sup>g</sup> | 5 |  | 18 |  |  |
| Income-to-Poverty ratio <sup>f</sup> |  |  |  |  |  |
| Below poverty level (<1.0) | 130 | 18 | 691 | 16 | 0.40 |
| Above poverty level (≥1.0) | 594 | 82 | 3562 | 84 |  |
| Missing <sup>g</sup> | 53 |  | 447 |  |  |
| Total 25 (OH) Vitamin D, nmol/L |  |  |  |  |  |
| < 30 | 49 | 7 | 329 | 8 | 0.80 |
| 30 – 74 | 422 | 60 | 2607 | 61 |  |
| ≥ 75 | 229 | 33 | 1329 | 31 |  |
| Mean ± SE <sup>d</sup> | 71 ± 1.14 |  | 69.8 ± 0.90 |  | <.0001 |
| Missing <sup>g</sup> | 77 |  | 435 |  |  |
| Tobacco, Pack-Years |  |  |  |  |  |
| 0 | 203 | 28 | 2530 | 56 | <.0001 |
| 1 – 10 | 122 | 17 | 787 | 18 |  |
| >10 | 409 | 55 | 1179 | 26 |  |
| Missing <sup>g</sup> | 43 |  | 204 |  |  |
| The six-month examination period |  |  |  |  |  |
| November 1 – April 30 | 307 | 40 | 2165 | 46 | 0.15 |
| May 1 – October 31 | 470 | 60 | 2535 | 54 |  |
| Spirometry Measurements (Mean ± SE <sup>d</sup>) |  |  |  |  |  |
| Baseline FEV1/FVC in % | 61.8±0.33 |  | 77.8±0.17 |  | <.0001 |
| Post-Bronchodilator FEV1 (% predicted) | 390 | 50 | 183 | 4 | <.0001 |
|  | 85.5±1.13 |  | 91.8±1.43 |  | <.0001 |
| Missing <sup>g</sup> | 387 |  | 4517 |  |  |

<sup>a</sup> Lower Limit of Normal
<sup>b</sup> Column Percentage from the total N. The percentage is calculated from the respective column based on Baseline FEV1/FVC
<sup>c</sup> Rao-Scott Chi-Square Test
<sup>d</sup> SE, standard error
<sup>e</sup> Body Mass Index (BMI)
<sup>f</sup> Income-to-Poverty ratio (\$Income/\$Threshold) = Represents the ratio of family or unrelated individual income to their appropriate poverty threshold. It was calculated by dividing family income by the poverty guidelines, specific to family size, appropriate year and state
<sup>g</sup> Missing values & Percent: represent column percent from the total N based on baseline FEV1/FVC.

### Study Participants’ Characteristics as related to the 25(OH)D Status

Those with vitamin D deficiency constituted only eight percent (*n*=569) of our sample (Table 2). In comparison to those with adequate serum vitamin D measurements, these participants had higher odds of being female gender(58% vs. 50.2%, *p*= 0.001), non-Hispanic Black race/ethnicity(52% vs. 15%, *p*=<.0001), obese with BMI ≥ 30 (50% vs. 39%, *p*= <.0001), and higher poverty-index-ratio < 1 (78% vs 82% *p*= <.0001), greater than 10 pack-year total lifetime smokers (38% vs. 31, *p*= 0.02), examined between November 1^st^ and April 30^th^ (62% vs 43%, *p*=<0001).

**Table 2.** Demographics and Clinical Characteristics of Study Participants by 25 (OH) Vitamin D Deficiency Status ^a^, NHANES, 2007-2010.

| Characteristic | 25 (OH) Vitamin D<br>≥ 30 (nmol/L) <sup>a</sup> |  | 25 (OH) Vitamin D<br>< 30 (nmol/L) <sup>a</sup> |  | p <sup>b</sup> |
| --- | --- | --- | --- | --- | --- |
|  | N | %<sup>c</sup> | N | %<sup>c</sup> |  |
|  | 6410 | 92 | 569 | 8 |  |
| Age, years |  |  |  |  |  |
| 40 – 49 | 1622 | 25 | 152 | 27 | <.0001 |
| 50 – 59 | 1453 | 23 | 150 | 26 |  |
| 60 – 69 | 1574 | 25 | 136 | 24 |  |
| ≥ 70 | 1761 | 27 | 131 | 23 |  |
| Mean ± SE <sup>d</sup> | 57.3 ± 0.24 |  | 57.5 ± 0.75 |  | <.0001 |
| Race |  |  |  |  |  |
| Mexican American | 1067 | 17 | 74 | 13 | <.0001 |
| Other Hispanic | 681 | 11 | 43 | 8 |  |
| Non-Hispanic White | 3465 | 54 | 132 | 23 |  |
| Non-Hispanic Black | 936 | 15 | 294 | 52 |  |
| Other race or Multi-racial | 259 | 4 | 26 | 5 |  |
| Gender |  |  |  |  |  |
| Male | 3195 | 49.8 | 239 | 42 | 0.001 |
| Female | 3215 | 50.2 | 330 | 58 |  |
| BMI <sup>e</sup>, kg/m<sup>2</sup> |  |  |  |  |  |
| < 18.5 | 83 | 1 | 14 | 2 | <.0001 |
| 18.5 – 25 | 1481 | 23 | 107 | 19 |  |
| 25 – 30 | 2340 | 37 | 156 | 28 |  |
| ≥ 30 | 2419 | 39 | 274 | 50 |  |
| Missing <sup>f</sup> | 87 |  | 18 |  |  |
| Income-to-Poverty ratio <sup>g</sup> |  |  |  |  |  |
| Below poverty level (<1.0) | 1043 | 18 | 114 | 22 | <.0001 |
| Above poverty level (≥1.0) | 4790 | 82 | 401 | 78 |  |
| Missing <sup>g</sup> | 577 |  | 54 |  |  |
| Tobacco, Pack-Years |  |  |  |  |  |
| 0 | 3255 | 53 | 251 | 47 | 0.02 |
| 1 – 10 | 979 | 16 | 85 | 15 |  |
| > 10 | 1877 | 31 | 204 | 38 |  |
| Missing <sup>g</sup> | 299 |  | 29 |  |  |
| The six-month examination period |  |  |  |  |  |
| November 1 – April 30 | 2766 | 43 | 350 | 62 | <.0001 |
| May 1 – October 31 | 3644 | 57 | 219 | 38 |  |

| **Spirometry Measurements (Mean ± SE <sup>d</sup>)** |  |  |  |
| --- | --- | --- | --- |
| Baseline FEV1(% Predicted) | 94.5 ± 0.43 | 89.5± 1.18 | <.0001 |
| Baseline FVC (% Predicted) | 97.6 ± 0.34 | 92.2± 1.10 | <.0001 |

<sup>a</sup> 30 nmol/L is the cutoff used to define vitamin D deficiency status
<sup>b</sup> Rao-Scott Chi-Square Test
<sup>c</sup> Column Percentage from the total N. The percentage is calculated from the respective column based on Baseline FEV1/FVC
<sup>d</sup> SE, standard error
<sup>e</sup> Body Mass Index (BMI)
<sup>f</sup> Missing values & Percent: represent column percent from the total N based on baseline FEV1/FVC.
<sup>g</sup> Income-to-Poverty ratio (\$Income/\$Threshold) = Represents the ratio of family or unrelated individual income to their appropriate poverty threshold. It was calculated by dividing family income by the poverty guidelines, specific to family size, as well as the appropriate year and state

### Association between Obstructive Lung Disease and Serum 25(OH)D

In addition to our primary predictor and outcome, our final model included race/ethnicity, BMI, pack-years smoking history, and poverty-index ratio. No statistically significant association was appreciated between obstructive lung disease and serum 25(OH)D in either the unadjusted or final model (Table 3). While interaction testing did not reveal a significant interaction between obstructive lung disease and self-reported race/ethnicity, there was a strong trend towards significance. To further explore this point, we performed repeated the final model, stratified by race/ethnicity. Results stratified by race/ethnicity revealed that, while not associated with non-Hispanic Whites, serum 25(OH)D levels were associated with obstructive lung disease in other (non-Mexican) Hispanics (OR [95%CI] = 1.48 [1.06-2.07], p=0.02) (Table 3). Among those with obstructive lung disease below the LLN, other (non-Mexican) Hispanics had a lower mean of serum 25(OH)D (65.4 nmol/L, 95% CI= 60.1-70.8), compared to non-Hispanic Whites (mean= 74.7, 95% CI= 72.0-77.4).

**Table 3.** The Crude and Multivariable-adjusted Associations between 25(OH)D and baseline FEV1/FVC<LLN ^a^, NHANES, 2007-2010.

| Vitamin D Measurements |  | Crude Model <sup>c</sup> |  |  | Final Model <sup>d</sup> |  |  |
| --- | --- | --- | --- | --- | --- | --- | --- |
|  |  | Odds Ratio | 95% CI. | *p*-value | Odds Ratio | 95% CI. | *p*-value |
| **25(OH)D**<br>Per 25 nmol/L <sup>b</sup> |  | 1.06 | 0.96-1.16 | 0.24 | 0.96 | 0.86-1.07 | 0.46 |
| **Stratified by Race** |  |  |  |  |  |  |  |
| **25(OH)D**<br>Per 25 nmol/L <sup>b</sup> | Mexican American | 1.19 | 0.90-1.58 | 0.21 | 0.91 | 0.70-1.19 | 0.48 |
|  | Other Hispanic | 1.46 | 1.05-2.02 | **0.03** | 1.48 | 1.06-2.07 | **0.02** |
|  | Non-Hispanic White (Ref.) | 0.98 | 0.88-1.10 | 0.77 | 0.94 | 0.83-1.06 | 0.29 |
|  | Non-Hispanic Black | 0.88 | 0.70-1.10 | 0.24 | 0.86 | 0.68-1.10 | 0.22 |
|  | Other race, including multi-racial | 1.33 | 0.91-1.96 | 0.14 | 1.57 | 0.84-2.93 | 0.16 |

<sup>a</sup> Lower Limit of Normal
<sup>b</sup> Total Vitamin D as continues variables, total Vitamin D= total Vitamin D/25, 1 unit change equal to a change by 25 nmol/L of vitamin D
<sup>c</sup> Crude model included only vitamin D measurement in the model.
<sup>d</sup> Final model included the following covariates: Vitamin D, Race, BMI, Smoking, and income-to-poverty ratio.

The mean of percentage predicted post-bronchodilator FEV1 was slightly higher in other (non-Mexican) Hispanics (mean= 89.8 %, 95% CI= 85.5-94.4), compared to non-Hispanic Whites (mean= 85.7, 95% CI=83.0-88.5). Additionally, the 95% Prediction Non-Mexican Hispanic’s ellipse is slightly thinner than other race subgroups, indicating that the correlation between baseline FEV1(% predicted) and total vitamin D is greater among Non-Mexican Hispanics (Figure 2).

**Figure 2.**
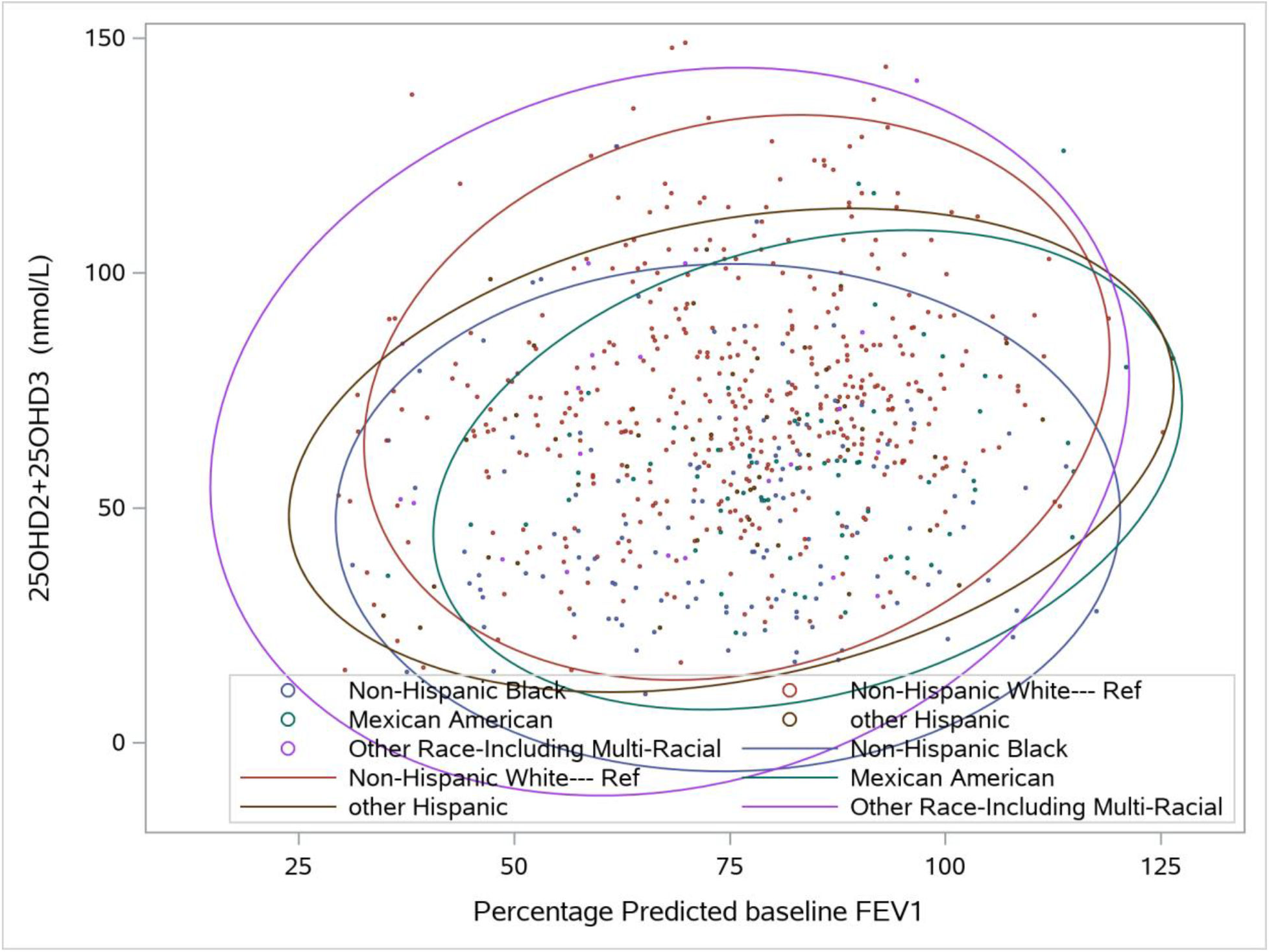
95% Prediction Ellipses for Baseline FEV1(% predicted) by each race subgroup among subjects with obstructive lung disease below the LLN. The means of the variables (the centers of the ellipses) are different across the race subgroups. The larger the ellipse, the greater the variance within that race subgroup. Abbreviations-FEV1: Forced Vital Capacity in the first second; LLN: lower limit of normal; 25(OH)D2+25(OH)D3: Total 25(OH)D.

## DISCUSSION

Using the NHANES 2007-2010 data, we explored an association between obstructive lung disease and serum 25(OH)D level in the general population. In our final model, vitamin D status was not independently associated with a diagnosis of obstructive lung disease. When stratified by race/ethnicity, we observed a positive association between serum Vitamin D levels and odds of obstructive lung disease among non-Mexican Hispanics.

The most similar study to our own, the population-based examination by Ganji et al., also failed to detect any association between vitamin D levels and obstructive lung disease.[28] In using ATS definitions rather than self-report, we addressed a major potential weakness of that study. By contrast, several previous studies have suggested such an association. For instance, a recent metanalysis found that as compared to those without COPD, those with the disease had a lower serum vitamin D level.[29] Notably, however, that report noted significant heterogeneity in the studies underlying that finding, and even in the meta-analysis had a smaller total sample size than reported in the present work. Further, while several studies report an association in COPD, this study examined the broader category of obstructive lung disease. The lack of association in other conditions may have diluted effects we could appreciate in a single disease state such as emphysema.

In light of studies suggesting a role for vitamin D in the pathogenesis of obstructive lung disease,[30,31] these results many appear counter-intuitive on facial analysis. However, lung development terminates in the third decade of life.[32] Evidence suggests that other vitamins influencing lung maturation, such as trans-retinoic acid, have a critical period for exerting some effects.[33] One might reasonably postulate a similar phenomenon impacting our results. That is, the key association with the risk of obstructive disease is not to present-day serum 25(OH)D levels, but those during some earlier phase of life. This distinction is especially important as vitamin D supplementation in all US populations increased considerably in the period immediately preceding—as well as throughout—this study.[34] Public health guidelines recommended increased intake in concert, and a great deal of work on the benefits of vitamin D in the lung was contemporaneous to this period. This timing raises the possibility that we have, in fact, captured the effect of deliberate intensification of therapy after the diagnosis of obstructive disease, an example of inverse correlation.

Unlike other ethnic groups, we did observe an association between serum hydroxy-vitamin D levels and obstructive lung disease among non-Mexican Hispanics. Several unknowns limit the interpretation of these results. It has previously been observed that low vitamin D levels in chronic respiratory disease may, in part, reflect reduced outdoor activity due to illness.[12] The multiple influences on 25(OH)D level and their ability to act as lurking variables is an important insight. Beyond debility from intrinsic disease, neighborhood quality may be an additional essential consideration. Hispanic-majority neighborhoods were burdened with higher environmental toxin exposure.[35,36] Further, there are positive correlations between air pollution exposure and walkability, especially in low-income communities.[37] These effects might plausibly account for higher serum 25(OH)D and higher incidence of obstructive lung disease, respectively. Alternatively, perhaps as a relatively low-cost health intervention, increased serum 25 (OH)D represents attempts to address problems from obstructive lung disease.

From a social determinant of health perspective, the resultant correlation might demonstrate the short-comings of over-reliance on individual-level interventions without addressing structural drivers of disease. However, Hispanics as a whole are under-served in healthcare and enormously under-represented in clinical trials.[38] From extant literature, Mexicans, compared to non-Mexican Hispanics, have been noted to have differences in likely immigration status, occupation type, and health insurance.[39] More fundamentally still, although NHANES data is structured as such, it is far from evident that a Cuban-American whose family was naturalized as refugees some three generations ago is meaningfully similar in circumstances or outlook to an undocumented Honduran migrant. One immediate implication of this heterogeneity can be seen in the cause of Puerto Ricans, who have a significantly higher prevalence of asthma than other Latino ethnicities[40]. Because they are one of the larger Latino groups within the US, any relationship between Vitamin D and obstructive lung disease in this group could create a false association for all non-Mexican Hispanics within our data set. It would be haphazard to offer in definitive explanation in a subset of an already under-studied and heterogeneous group. While differing determinants of Vitamin D status might explain its varying association with obstructive lung disease, the paucity of evidence at this time precludes any conclusion. These results require confirmation and then further exploration through dedicated and well-designed studies in the target population.

The present work does not preclude other vital roles for vitamin D in confronting public health concerns related to obstructive lung disease. Our results only examined the association between current serum 25-hydroxy vitamin D levels and the diagnostic criteria for obstructive lung disease. It may, for instance, influence the degree of obstruction once present. Other investigative teams report exactly these findings.[28] Symptom burden and exacerbation also have poor correlation with measured obstruction.[20] vitamin D levels might, therefore, correlate with disease control independent of its relationship to measured severity. Though limited by their small sample size, some randomized controlled trials have reported improvements in exacerbations, 6-minute walk distance, or symptoms as measured by the COPD Assessment Test.[41,42] Longitudinal cohorts of COPD populations similarly demonstrate increased exacerbations, accelerated FEV1 decline, and higher symptom burden in association with lower vitamin D levels.[43]

Our study had several other important limitations. The cross-sectional design precludes any discussion of causality. While skin complexion affects vitamin D absorption[44], and African ancestry is associated with lower mean lung function,[45] self-identified race can be a poor proxy for both.[46,47] Co-morbid restrictive disease can obscure underlying obstructive physiology, but the NHANES omitted body plethysmography that would allow for its assessment. While we observed essential effects related to socioeconomic, this was measured in only one dimension at the individual level and not at all at the neighborhood level, limiting our appreciation of its effects.

In a population-based study of the United States derived from the NHANES 2007-2008 and 2009-2010 cycles, in the general population, we found no association between serum 25(OH)D levels and a diagnosis of obstructive lung disease. These findings may have been impacted by increasing public awareness about the physiologic benefits of vitamin D, or the exclusion of those with more severe obstructive disease. Among non-Mexican Hispanics, we found increasing serum 25(OH)D levels had increased odds for obstructive lung disease. This finding calls attention to the importance of further research on minority health as the United States grows increasingly diverse. Altering the prevalence of obstructive lung disease in the United States may require interventions that are more comprehensive, longitudinal, or both, to yield substantive improvement.

## Data Availability

The National Health and Nutrition Examination Survey (NHANES) is an ongoing national survey project of the Center for Disease Control and Prevention (CDC) designed to assess the general US population's health. Also, NHANES is a major program of the National Center for Health Statistics (NCHS), which is part of the CDC and responsible for producing vital and health statistics for the Nation.
The survey is unique in that it combines interviews and physical examinations. Data are collected annually on a 2-year cycle, using a multistage, probability-sampling design to generate population-level estimates. In our study, we included all participants from the 2007-2008 and 2009-2010 survey cycles.
Information and data are made available, on the NHANES website, to the public and researchers worldwide.

https://www.cdc.gov/nchs/nhanes/index.htm

https://www.cdc.gov/nchs/data/nhanes/nhanes_release_policy.pdf

https://www.n.cdc.gov/nchs/nhanes/continuousnhanes/default.aspx?BeginYear=2007

https://www.n.cdc.gov/nchs/nhanes/ContinuousNhanes/Default.aspx?BeginYear=2009

## Acknowledgments

The authors would like to thank the faculty of the Rollins School of Public Health at Emory University, more specifically, the thesis committee chair, Veronika Fedirko, MPH, Ph.D., for her tremendous guidance and support with the implementation of the algorithm, study design, data cleaning, and analysis. Additionally, the authors would like to thank Dr. Mehrdad Arjomandi, Dr. Prescott Woodruff, and Dr. Neeta Thakur for their valuable review of the manuscript.

